# Diagnostic value of neutrophil gelatinase-associated lipocalin and neutrophil-to-lymphocyte ratio in type 2 diabetes mellitus individuals with reduced estimated glomerular filtration rate(eGFR): A case-control study in a municipal hospital in Ghana

**DOI:** 10.1101/2024.10.31.24316479

**Authors:** Allwell Adofo Ayirebi, Wina Ivy Ofori Boadu, Benedict Sackey, Lilian Antwi-Boateng, Prince Adoba, Patrick Adu, Joseph Boachie, Stephen Twumasi, Joseph Frimpong, Emmanuel Ekow Korsah, Ezekial Ansah, Ebenezer Senu, Eugene Arele Ansah, Daniel Nii Martey Antonio, Afua Marfowaa, Isaac Opoku Antwi, John Agyemang Sah, Joachim Baba Domosie, Abiba Khalifah, Evans Adu Asamoah, Christian Obirikorang, Otchere Addai-Mensah, Enoch Odame Anto

**Author notes:** **Correspondence:** Allwell Adofo Ayirebi, Enoch Odame Anto ( _.

## Abstract

**Introduction:** Serum neutrophil gelatinase-associated lipocalin (sNGAL), a renal tubular marker, and Neutrophil-to-Lymphocyte Ratio (NLR), a haematological inflammatory marker are two biomarkers that have recently received attention, because of their association with kidney disease. This study examined the diagnostic value sNGAL and NLR in type 2 diabetes mellitus (T2DM) patients with reduced eGFR.

**Materials and methods:** In this hospital-based case-control research, 97 T2DM participants and 70 healthy subjects were included. Participants’ information was documented using a structured questionnaire and patient case records. Venous blood was drawn from each participant to evaluate absolute neutrophil and lymphocyte count ratio, glycosylated haemoglobin (HBA1c), creatinine, sNGAL, and fasting blood glucose.

**Results:** sNGAL and NLR were higher in diabetes patients with reduced eGFR than those with normal eGFR and control (p < 0.05). sNGAL level was negative correlated with eGFR among both good (r= -0.317, *p*= 0.036) and poor (r= -0.544, *p*< 0.001) glycaemic-controlled T2DM subjects with reduced eGFR. A negative correlation was observed between NLR and eGFR among poor glycaemic-controlled T2DM subjects with reduced eGFR (r= -0.329, *p*= 0.016). At a cut off of 8.87 µg/L and 2.34 respectively, sNGAL and NLR were found to be good predictors of nephropathy among T2DM patients (AUC=100.0, p< 0.0001 and AUC=76.0, p< 0.0001 respectively) with sNGAL being the superior marker.

**Conclusions:** sNGAL and NLR have important diagnostic value for diabetic with reduced eGFR. Whiles sNGAL showed superiority and is recommended, NLR can serve as a less expensive and readily measurable alternative biomarker for diabetic patients with reduced eGFR, particularly in poorly controlled diabetes. These two markers can be added to the available array of tests used to indicate nephropathy in type 2 diabetics to help clinicians in better management of the disease.

## Introduction

Type 2 diabetes mellitus (T2DM) is a metabolic illness that affects most people worldwide. Globally, the prevalence of diabetic mellitus (DM) is believed to be at 8.5%, and it is predicted that this number could double in the next ten years (1). Numerous research has indicated that the prevalence of diabetes mellitus in Ghana varies from 6.4% to 13.9% (2,3).

Diabetic Nephropathy (DN), as a major vascular adverse outcome of diabetes mellitus (DM), arises due to uncontrolled hyperglycaemia resulting in glomerular and tubular destruction and eventual permanent kidney damage (4). A lower estimated glomerular filtration rate (GFR) of less than 60 ml/min/1.73 m2 and albuminuria for a minimum of three months are indicative of chronic kidney disease (5). The prevalence of diabetic nephropathy in Ghana ranges from 14% to 19% (6). A 13-year retrospective study found diabetic nephropathy to be the third leading cause of chronic kidney disease, and the second leading cause of end-stage kidney disease in Ghanaian adults (7).

The conventional estimated Glomerular Filtration Rate (eGFR) and its albuminuria correlation (uACR) are regarded as the primary laboratory markers for indicating chronic kidney disease (CKD) in Ghana. However, creatinine is a marker of glomerular filtration and is unable to appropriately assess renal tissue damage. Also, serum creatinine rises after about 50% of kidney function is lost, hence not sensitive enough to pick up on subtle variations in renal function that might occur before albuminuria manifests. (8,9). It has been widely proposed that other biomarkers are needed for early detection of renal dysfunction. There is also the need for cost-effective and accessible biomarkers that can effectively detect DN, as current markers are expensive and financially burdensome for both the government and individuals (10).

A tubulointerstitial injury marker, neutrophil gelatinase-associated lipocalin (NGAL), is a 25kDa protein that stems from the lipocalin superfamily has been suggested as a potential marker of CKD (11). It is mainly used to transport tiny hydrophobic molecules and control innate immune responses. It is secreted by renal tubular epithelial cells and released by active neutrophils (12). The haematological inflammatory neutrophil-to-lymphocyte ratio (NLR) measures the interaction between the innate and adaptive cellular immune responses. NLR is an affordable and easily measurable marker of inflammation, indicating inflammatory burden in many chronic conditions. It has been shown that NLR can serve as an early marker for diabetic nephropathy (DN), evidenced by a lower predicted risk of hospitalizations in diabetic patients undergoing haemodialysis (13). The efficacy of these two markers in detecting DN has not been evaluated in the context of Ghana. As a result, the study assessed the performance of sNGAL and NLR in predicting nephropathy among type 2 diabetics. Moreover, given the major role uncontrolled hyperglycaemia plays in renal pathology incidence, this study also assessed these markers in diabetics with reduced eGFR with good and poor glycaemic management.

## Materials and Methods

### Study design, duration and study setting

This was a prospective case-control study and conducted at the Ejisu Government Hospital from December 15,2022 to February 25, 2023.The Ejisu Government Hospital, located in the Ejisu Juaben Municipality of the Ashanti region of Ghana, served as the site of this hospital-based case-control study. The municipality has a total size of 637.2 square kilometres, or about 10% of the Ashanti Region’s total land area. The geographical coordinates of the location in question are situated between longitudes 1°5W and 1°39W, and latitudes 7°9’ N and 7°36°N. The hospital is the biggest healthcare facility in the municipality, providing an extensive array of services including emergency medical treatment, obstetrics and gynaecology, laboratory testing, and medical imaging. The medical centre is open around-the-clock, all year round. Participants were chosen for the study from the facility’s diabetic clinic.

### Ethical Clearance and Informed Consent

The Committee for Human Research Publication and Ethics of Kwame Nkrumah University of Science and Technology, School of Medical Sciences, evaluated and approved the study protocol, permission forms, and participant information material (CHRPE/AP/742/22). The Ejisu Government Hospital ethical committee also approved the study. Once the study participants had received a thorough explanation of the study’s objectives, advantages, risks, and right to withdraw at any time in both English and the local dialect (primarily Twi), their written consent was requested.

### Study participants

A total of 167 participants were recruited for this study. Cases consisted of 97 type 2 diabetics (21 males and 75 females) under management at the clinic, six months or more prior to recruitment of study, and 70 participants without diabetes (28 men and 42 females) used as healthy controls. Patients with cancer, microbial infections, pregnant women, those with changes in leucocyte count, and advanced kidney impairment (eGFR <30 ml/min) were excluded. Additionally, we excluded diabetics with iron deficiency anaemia, HIV infection, hepatitis C, and inherited haemolytic disorders.

### Data collection

Demographic and clinical history of the participants were recorded using well-structured questionnaires and patient case records. Dietary habits were categorised qualitatively as occurring rarely (one per month), irregularly (seven times per month), and regularly (twice per month). Estimates were made for height to the nearest centimetre without shoes and weight to the nearest 0.1 kg when wearing light clothes. A wall-mounted ruler was used to measure height. A Zhongshan Camry Electronic Co. Ltd. bathroom scale (Guangdong, China) was used to assess weight.

### Blood sampling

After an 8-12 hours overnight fast, venous blood sample was drawn from each participant into fluoride/oxalate, serum separator tube (SST) and ethylenediamine tetracetate (EDTA) tubes. After clotting, the SST was centrifuged and kept at -20°C until analysis.

### Laboratory Assays

Haemogram analysis of the EDTA samples were determined within one (1) hour after sample using an automated haematology analyser (XN-550; Sysmex Corporation, Kobe Japan). NLR levels were determined from haemogram test by calculating the ratio of absolute neutrophil count and absolute lymphocyte count. Glycosylated haemoglobin test was run on a Getein Biotech Immunofluorescence Quantitative Analyser (Getein 1100, China). Well controlled diabetes and poorly controlled diabetes was defined as HbA1c ≤7% and >7% respectively. The Mindray BS-240 fully automated chemistry analyser and an assay kit (Shenzhen Mindray Bio-Medical Electronics Co., China) were used to estimate the plasma concentration and level of creatinine in serum samples. Estimated Glomerular Filtration Rate (eGFR) was calculated using the CKD-EPI equation (16). Participants with eGFR <60ml/min/1.72m^2^ were classified as having reduced eGFR. A commercially available ELISA kit (Melson Shanghai Chemical Ltd, China) was used for the sNGAL measurement. Reagent was used in accordance with the manufacturer’s instructions to measure samples from the controls and the participants using the solid phase ELISA technique.

### Statistical analysis

The data obtained from the study was entered into Microsoft Excel software (2016) and analysed using GraphPad Prism (version 8.0.1), R programming version 4.2.3, and IBM Statistical Package for Social Sciences (version 26.0). In contrast to categorical variables, which were expressed as proportions and compared using the chi-square test, parametric continuous variables were expressed as mean standard deviation. The chi-square test was used to examine the relationship between categorical research variables and diabetics with reduced eGFR. Additionally, one-way ANOVA and the post-hoc Bonferroni test were used to assess the differences between parametric continuous variables and study groups. Pearson correlation was used to analyse the association between NGAL, NLR, and eGFR. The area under the curves for NGAL and NLR used in the evaluation of diabetic nephropathy was calculated using the Receiver Operator Characteristics Curve. A p value of 0.05 and a 95% confidence interval were used for all statistical significance calculations.

## Results

### Baseline characteristics of study participants

The prevalence of reduced eGFR among type 2 diabetic was 10.4%. The majority of the case group participants that enrolled for the study were females, who had no postsecondary education, and had an average age of 57 years. Body mass index, fasting blood glucose, diastolic and systolic blood pressure, glycosylated haemoglobin percent, and creatinine levels, showed statistically significant difference (p<0.05) among diabetics with reduced eGFR, those with normal eGFR, and healthy controls. The variables among diabetics with reduced eGFR and diabetics with normal eGFR were considerably greater than those among healthy controls in a post-hoc Bonferroni test (p<0.05), with the exception of diastolic blood pressure. White blood cell count, absolute neutrophil count, absolute lymphocyte count and NLR were higher diabetes patients with reduced eGFR than those with normal eGFR and controls (p < 0.05), but were comparable between controls and diabetes patients with normal eGFR (p > 0.05). Similarly, sNGAL levels were increased in diabetes patients with redced eGFR than those with normal eGFR and controls (p < 0.05). A significant association was found between consumption of sweets, vigorous exercising and development of diabetics with reduced eGFR among study participants (p<0.05) Conversely, alcohol intake was not significantly associated with reduced eGFR among study participants (*p*>0.05) **(Table 1).**

**Table 1.**
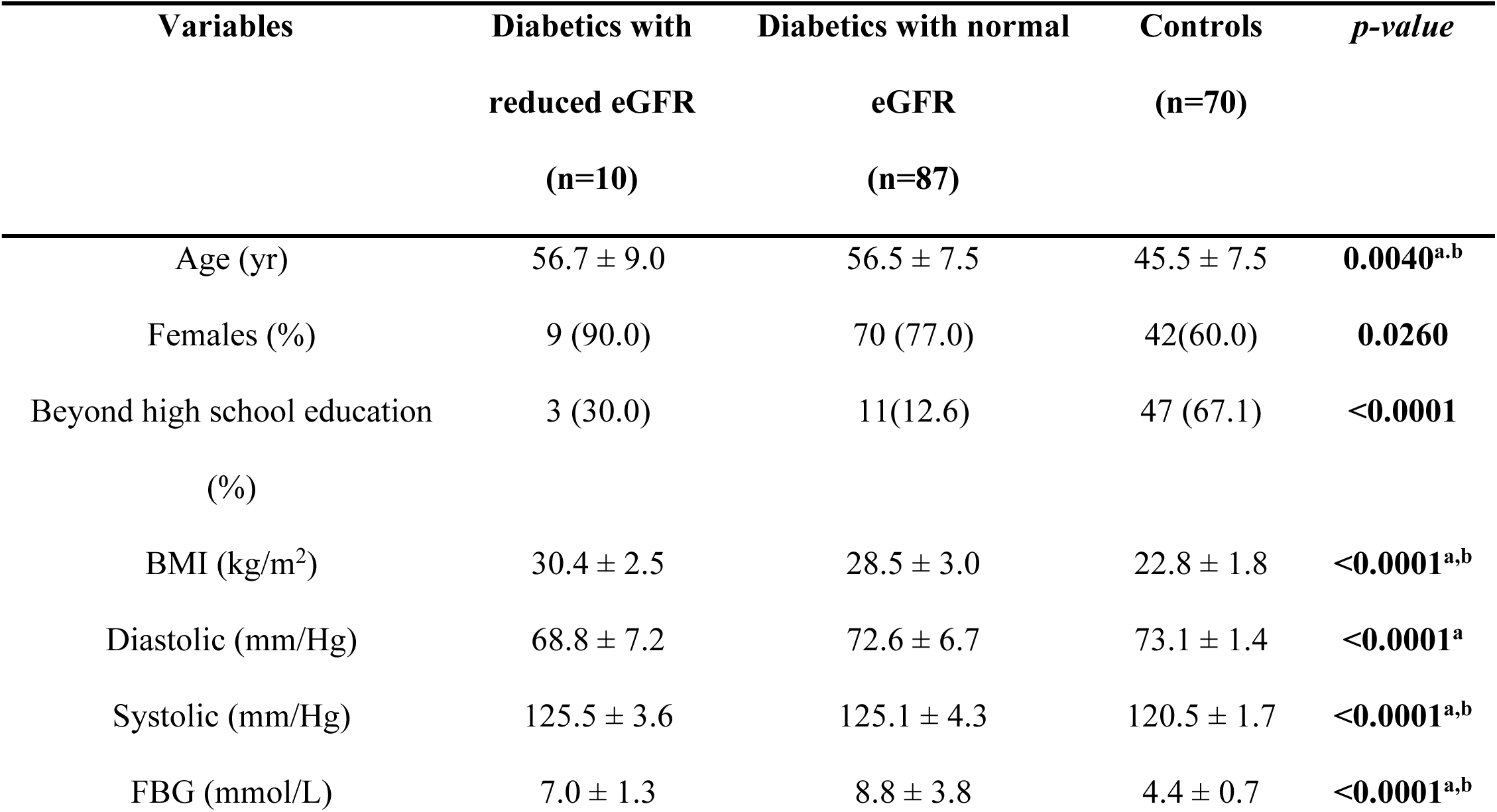
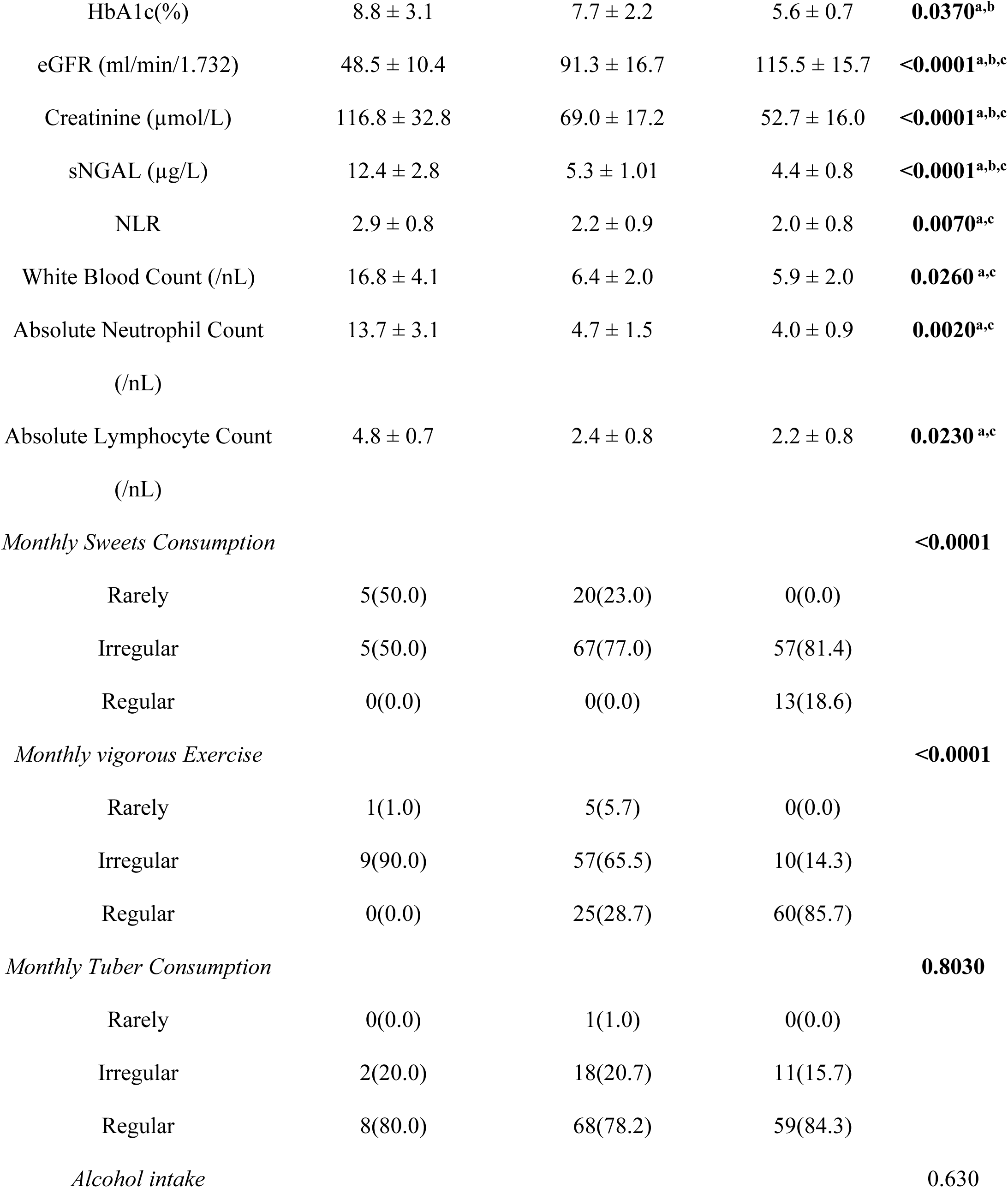
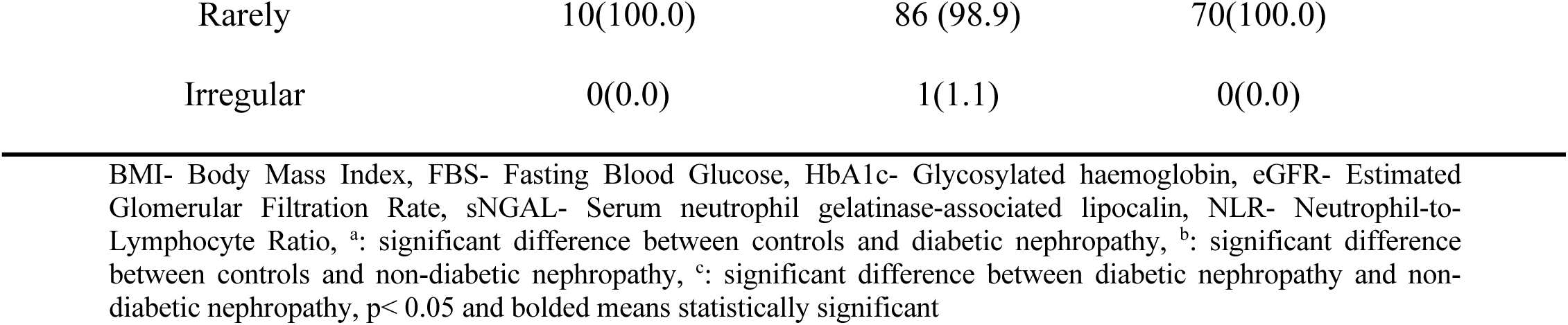
Comparison of sociodemographic, anthropometric indices, haemato-biochemical values and dietary lifestyle of type 2 diabetics with reduced eGFR and normal eGFR and controls.

| Variables | Diabetics with<br>reduced eGFR<br>(n=10) | Diabetics with normal<br>eGFR<br>(n=87) | Controls<br>(n=70) | *p-value* |
| --- | --- | --- | --- | --- |
| Age (yr) | 56.7 ± 9.0 | 56.5 ± 7.5 | 45.5 ± 7.5 | **0.0040<sup>a,b</sup>** |
| Females (%) | 9 (90.0) | 70 (77.0) | 42(60.0) | **0.0260** |
| Beyond high school education<br>(%) | 3 (30.0) | 11(12.6) | 47 (67.1) | **<0.0001** |
| BMI (kg/m<sup>2</sup>) | 30.4 ± 2.5 | 28.5 ± 3.0 | 22.8 ± 1.8 | **<0.0001<sup>a,b</sup>** |
| Diastolic (mm/Hg) | 68.8 ± 7.2 | 72.6 ± 6.7 | 73.1 ± 1.4 | **<0.0001<sup>a</sup>** |
| Systolic (mm/Hg) | 125.5 ± 3.6 | 125.1 ± 4.3 | 120.5 ± 1.7 | **<0.0001<sup>a,b</sup>** |
| FBG (mmol/L) | 7.0 ± 1.3 | 8.8 ± 3.8 | 4.4 ± 0.7 | **<0.0001<sup>a,b</sup>** |
| HbA1c(%) | 8.8 ± 3.1 | 7.7 ± 2.2 | 5.6 ± 0.7 | **0.0370<sup>a,b</sup>** |
| eGFR (ml/min/1.732) | 48.5 ± 10.4 | 91.3 ± 16.7 | 115.5 ± 15.7 | **<0.0001<sup>a,b,c</sup>** |
| Creatinine (μmol/L) | 116.8 ± 32.8 | 69.0 ± 17.2 | 52.7 ± 16.0 | **<0.0001<sup>a,b,c</sup>** |
| sNGAL (μg/L) | 12.4 ± 2.8 | 5.3 ± 1.01 | 4.4 ± 0.8 | **<0.0001<sup>a,b,c</sup>** |
| NLR | 2.9 ± 0.8 | 2.2 ± 0.9 | 2.0 ± 0.8 | **0.0070<sup>a,c</sup>** |
| White Blood Count (/nL) | 16.8 ± 4.1 | 6.4 ± 2.0 | 5.9 ± 2.0 | **0.0260<sup>a,c</sup>** |
| Absolute Neutrophil Count<br>(/nL) | 13.7 ± 3.1 | 4.7 ± 1.5 | 4.0 ± 0.9 | **0.0020<sup>a,c</sup>** |
| Absolute Lymphocyte Count<br>(/nL) | 4.8 ± 0.7 | 2.4 ± 0.8 | 2.2 ± 0.8 | **0.0230<sup>a,c</sup>** |
| *Monthly Sweets Consumption* |  |  |  | **<0.0001** |
| Rarely | 5(50.0) | 20(23.0) | 0(0.0) |  |
| Irregular | 5(50.0) | 67(77.0) | 57(81.4) |  |
| Regular | 0(0.0) | 0(0.0) | 13(18.6) |  |
| *Monthly vigorous Exercise* |  |  |  | **<0.0001** |
| Rarely | 1(1.0) | 5(5.7) | 0(0.0) |  |
| Irregular | 9(90.0) | 57(65.5) | 10(14.3) |  |
| Regular | 0(0.0) | 25(28.7) | 60(85.7) |  |
| *Monthly Tuber Consumption* |  |  |  | **0.8030** |
| Rarely | 0(0.0) | 1(1.0) | 0(0.0) |  |
| Irregular | 2(20.0) | 18(20.7) | 11(15.7) |  |
| Regular | 8(80.0) | 68(78.2) | 59(84.3) |  |
| *Alcohol intake* |  |  |  | **0.630** |

|  |  |  |  |
| --- | --- | --- | --- |
| Rarely | 10(100.0) | 86 (98.9) | 70(100.0) |
| Irregular | 0(0.0) | 1(1.1) | 0(0.0) |

BMI- Body Mass Index, FBS- Fasting Blood Glucose, HbA1c- Glycosylated haemoglobin, eGFR- Estimated Glomerular Filtration Rate, sNGAL- Serum neutrophil gelatinase-associated lipocalin, NLR- Neutrophil-to- Lymphocyte Ratio, <sup>a</sup>: significant difference between controls and diabetic nephropathy, <sup>b</sup>: significant difference between controls and non-diabetic nephropathy, <sup>c</sup>: significant difference between diabetic nephropathy and non- diabetic nephropathy, p< 0.05 and bolded means statistically significant

### The distribution and comparison of sNGAL and NLR between healthy controls, and good and poor glycaemic controlled diabetics

We observed a significantly higher NLR in poor glycaemic controlled diabetics with reduce eGFR than good glycaemic control and healthy subjects [2.09 (1.73 – 2.78) vs 1.81(1.36 – 2.22) vs 1.72 (1.40 – 2.51) respectively, (*p*<0.05)]. sNGAL level was also significantly higher among diabetics with reduced eGFR patients with poor and good glycaemic control than the healthy individuals [{5.60 (4.8 – 6.20) vs 5.18 (4.88 – 6.11) vs 4.50 (3.60 – 5.10)} µg/L respectively, (*p*<0.05)]. NLR levels between good glycaemic control and controls as well as sNGAL levels between poor and good glycaemic controlled patients with reduced eGFR were not statistically significant (*p*>0.05)**(Figure 1).**

**Figure 1:**
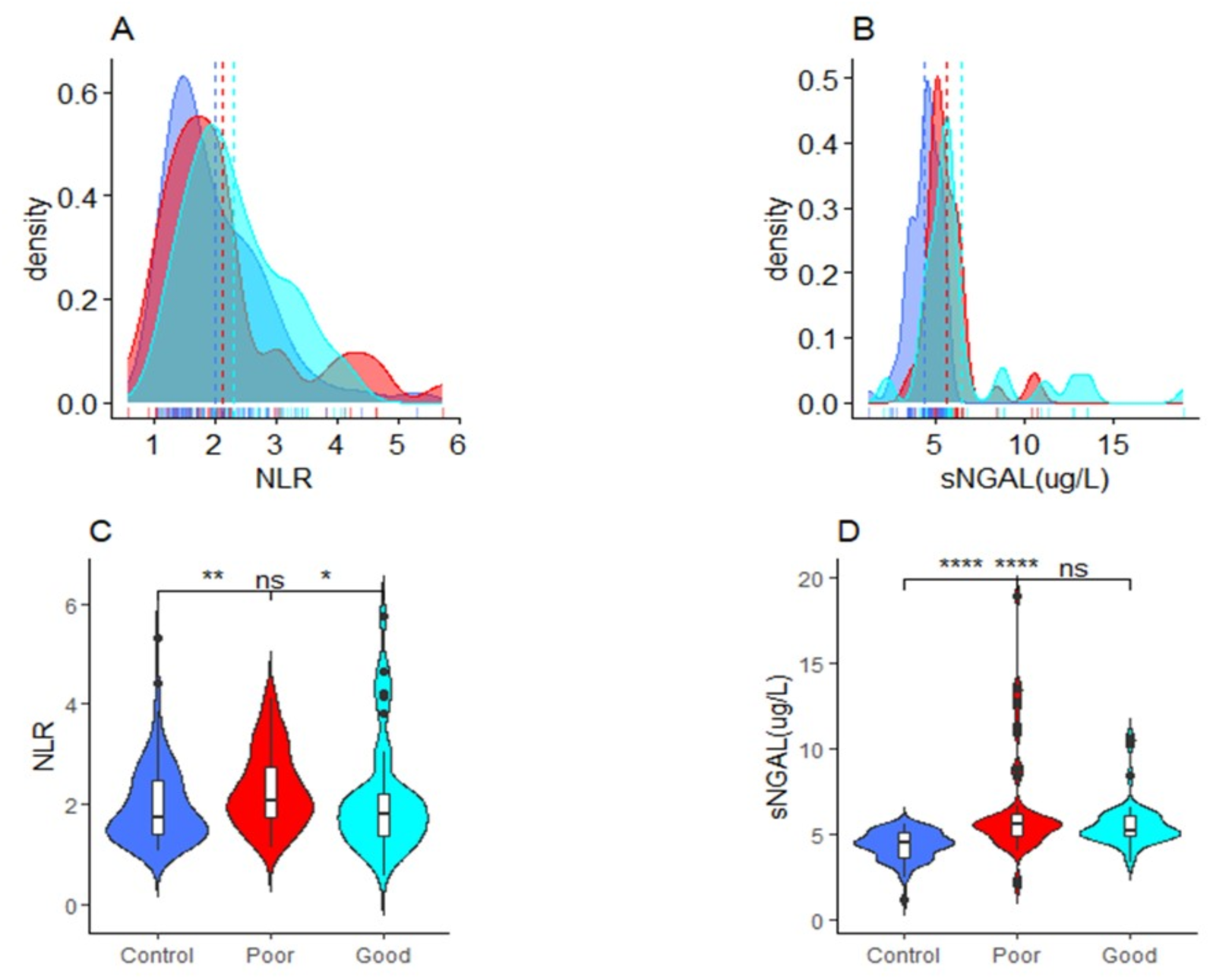
Density (A-B) and box and violin plot (C-D) showing the distribution and comparison respectively of NLR, and sNGAL between good and poor glycaemic controlled type 2 diabetics with reduced eGFR and healthy controls; *ns (not significant) = p > 0.05, * = p < 0.05, ** = p < 0.01, *** = p < 0.001, **** = p < 0.0001*

### Pearson correlations between sNGAL, NLR and eGFR in good and poor glycaemic controlled diabetics with reduced eGFR

We found an insignificant negative weak correlation between NLR and eGFR (r = -0.038, *p* = 0.806) **(Figure 2A)** and a significant moderate negative correlation between sNGAL and eGFR (r = -0.317, *p* = 0.036) (**Figure 2B)** among good glycaemic controlled diabetics with reduced eGFR. There were also significant weak negative correlations between NLR and eGFR (r = -0.329, *p* = 0.016) **(Figure 2C)** and a significant moderate negative correlation between sNGAL and eGFR (r = -0.544, *p* < 0.001) (**Figure 2D)** among poor glycaemic controlled diabetics with reduced eGFR.

**Figure 2:**
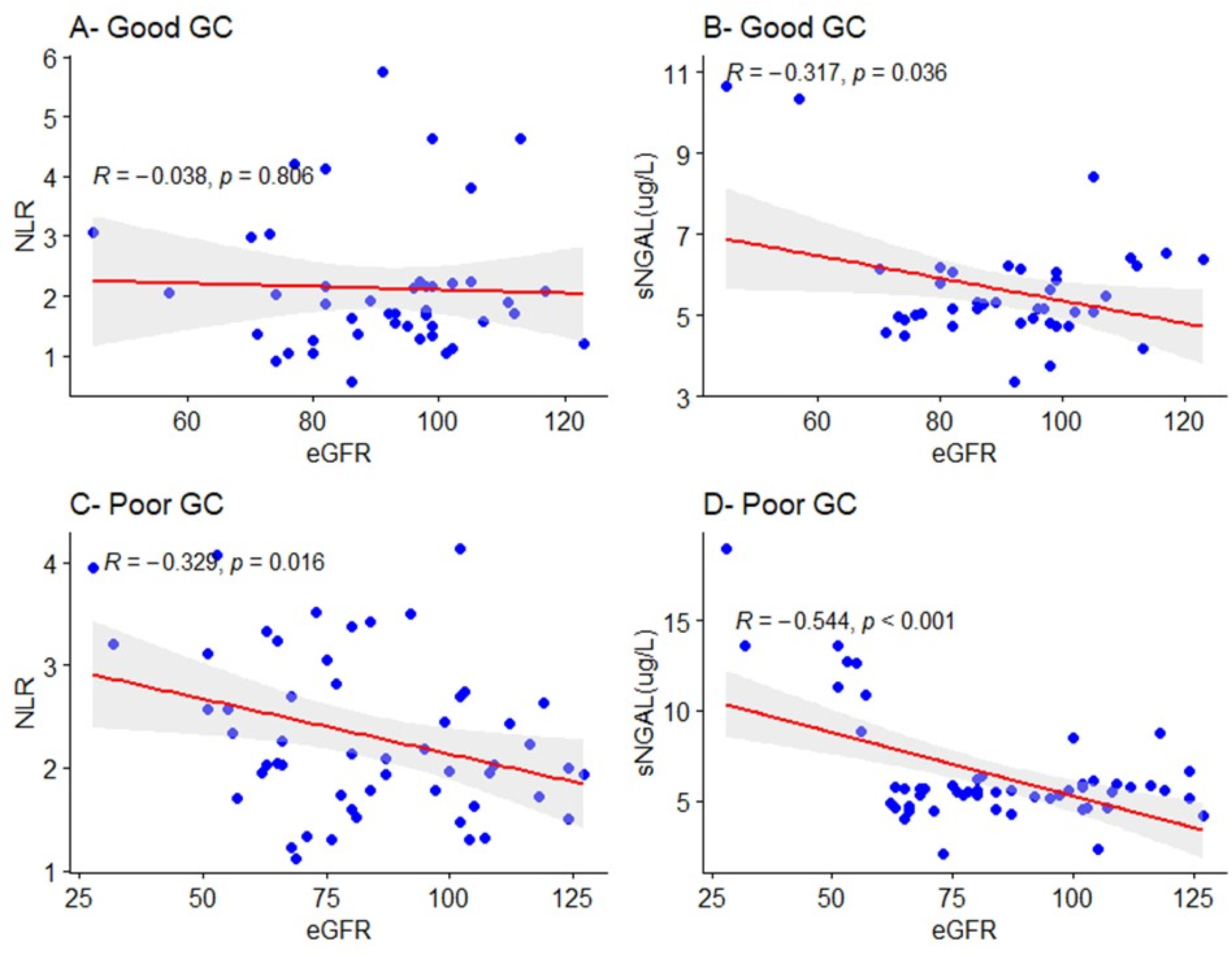
Relationship between sNGAL NLR, and eGFR among good and poor glycaemic controlled type 2 diabetics with reduced eGFR.

### Diagnostic Performance of NGAL and NLR

Receiver operating characteristic (ROC) analysis showed that sNGAL had an excellent performance (AUC = 100.0, *p* < 0.0001) and NLR had a moderate performance (AUC = 76.0, *p* < 0.0001) for predicting diabetic nephropathy **(Figure 3).** An NGAL cut-off of 8.87µg/L had 100% sensitivity and specificity while NLR threshold of 2.34 had 80% sensitivity and 73.6% specificity in predicting nephropathy in type 2 diabetes mellitus patients using CKD-EPI equation.**(Table 2).**

**Figure 3:**
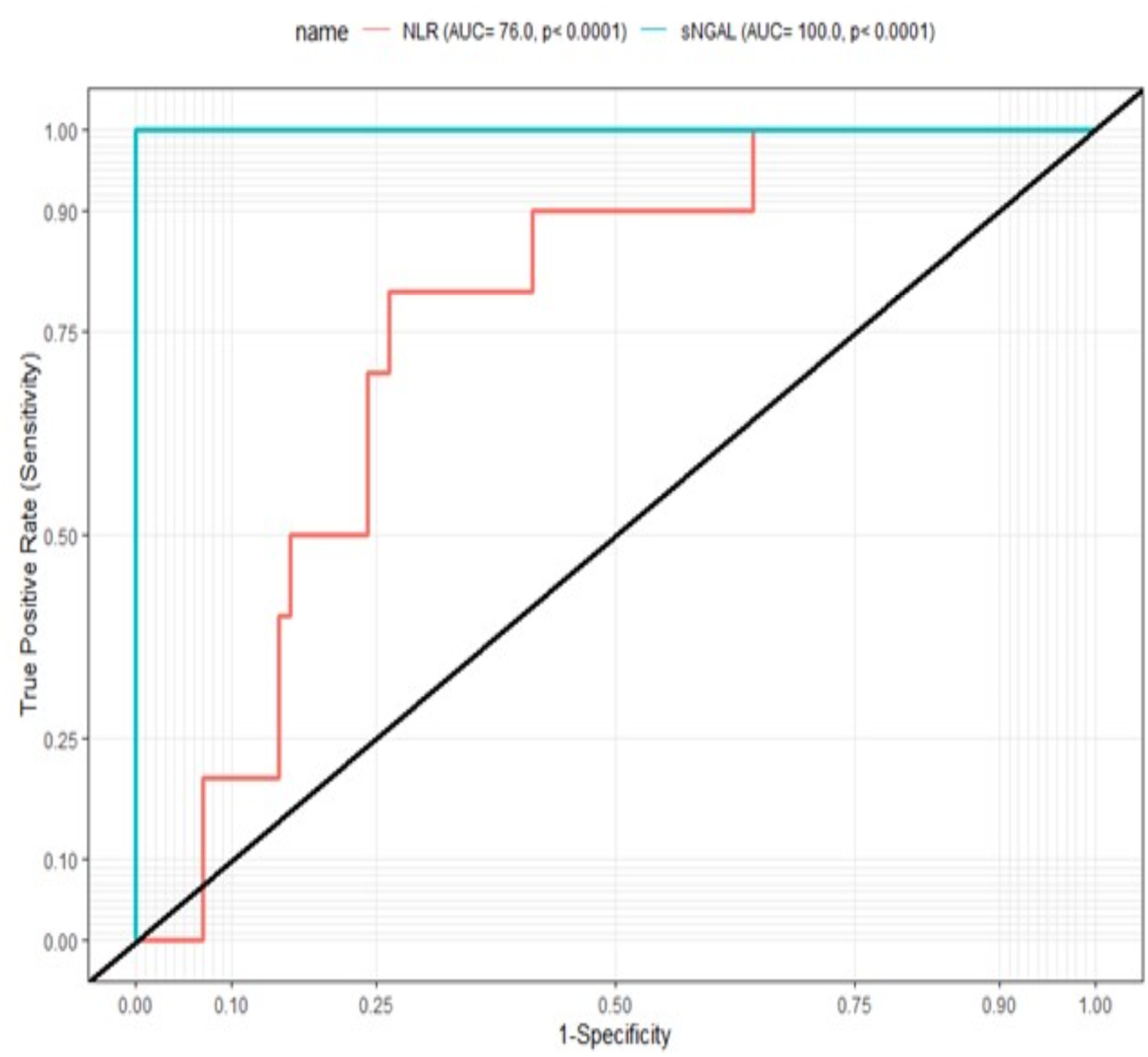
The receiver operating characteristics (ROC) curves of NGAL and NLR for predicting diabetic nephropathy among T2DM.

**Table 2.** Diagnostic performance of sNGAL and NLR in predicting Diabetic Nephropathy among type 2 diabetics.

| Marker | Cut-off | Sensitivity (95% CI) | Specificity (95% CI) | PPV | NPV | LR+ | LR- |
| --- | --- | --- | --- | --- | --- | --- | --- |
| NLR | 2.34 | 80.0 (47.8-95.1) | 73.6 (63.4-81.7) | 25.8 | 97 | 3.03 | 0.27 |
| sNGAL (µg/L) | 8.87 | 100.0 (67.4-100.0) | 100.0 (94.8-100.0) | 100.0 | 100.0 | inf | 0.00 |

Abbreviations: NLR: Neutrophil-to-Lymphocyte Ratio, sNGAL: serum Neutrophil Gelatinase – Associated Lipocalin, CI: Confidence interval, PPV: Positive predictive value, NPV: Negative predictive value, LR+: Positive likelihood ratio, LR–: Negative likelihood ratio

## Discussion

Effective monitoring is paramount to preventing exacerbating of diabetes mellitus to diabetes nephropathy (DN). This can be achieved by identifying biomarkers that can provide early indication of renal impairment as well as markers that are readily available and less expensive. This hospital–based case-control study aimed to investigate the diagnostic value of sNGAL and NLR for the detection of nephropathy among type 2 diabetics using the CKD-EPI equation. The prevalence of reduced eGFR among T2DM patients from this present study was significant (10.4%) and reflective of findings by Tannor et al., and Ofori et al, (6,15) which reported between them 16.1% to 19% prevalence of CKD based on reduced eGFR among diabetes mellitus patients in Ghana. The variations of prevalences may be due to the different samples sized. Nevertheless, the prevalence observed in this is alarming and could be attributed to patients not taking their medications as directed by physicians or the therapy regimen not working for them.

In this study, NLR level was significantly higher in diabetics with redduced eGFR than those with normal eGFR, consistent with findings from Paulus et al, (17) who reported an increase in NLR levels among diabetics with reduced eGFR than those with normal eGFR. This could be attributed to excess glucose in circulation causing damage to the renal tubules, subsequently inducing a persistent inflammatory state. However, studies conducted by Wan et al, (18) reported comparable levels of NLR in diabetics with reduced eGFR and with normal eGFR. The reason for this inconsistency includes differences in sample size, population and geography. In addition, a significant increase in NLR was found in diabetics with reduced eGFR with poor glycaemic control than those with good glycaemic control and is reflective of the findings from Hussain et al., (19) that suggested that an elevated HbA1c is associated with high NLR levels due to an induced chronic inflammatory state by the hyperglycaemic milieu.

sNGAL levels in this study was found to be significantly higher in diabetics with reduced eGFR than those with normal eGFR. This could be due to damage to renal tubules as a result of the presence of excess glucose in circulation leading to endothelial dysfunction and tubulointerstitial damage consistent with finding from Kaul et al, (20). Furthermore, mild rise in levels of serum sNGAL level was also found in diabetics with reduced eGFR with both poor and good glycaemic control than apparently healthy non-diabetics which also agrees with Kaul et al., (20) that sNGAL is an early predictor of DN in type 2 diabetics. This explains why even in good glycaemic controlled diabetics, levels of sNGAL were mildly increased.

Moreover, sNGAL level in both good and poor glycaemic controlled diabetic patients was negatively associated with eGFR. This is consistent with the findings from Bolignano, Lacquaniti, et al., which suggested an inverse correlation between NGAL and eGFR, thus proposing NGAL as a strong and independent risk marker for progression of chronic kidney disease (21). It is also in line with the findings from Danquah et al, (22) which reported that plasma NGAL was a reliable indicator of kidney damage in initial phases of CKD in comparison to serum creatinine. This indeed makes sNGAL a good marker for kidney injury as even before glycaemic control worsens, levels are already elevated mildly or markedly.

We also found that NLR and eGFR among poorly glycaemic controlled diabetic patients was negatively associated. However, no such correlation was observed in good glycaemic controlled diabetes patients. The poor management of hyperglycaemia leads to damage of glomerulus and tubules of nephron and may results in neutrophilia caused by release of cytokines and chemokines. This study also examined the diagnostic performance of these markers as indicators of DN. The excellent cut point to indicate DN was assessed using CKD-EPI equation as reference. This study found NLR (AUC = 76.0) and NGAL µg/L (AUC = 100) to be good indicators of DN with sNGAL being the superior marker. These findings is consistent with previous studies which observed good performance of these markers for diagnosing DN (18,22–24). Also, in this present study, the optimal threshold at which NLR and NGAL levels indicated DN were 2.34 and 8.87 µg/L respectively. However, the cut-off level for sNGAL were markedly lower than reported findings of Kaul et al, (20) with cut point of 78.7ng/ml (sensitivity 95.1% ; specificity 100%) and Danquah et al, (22) with cut point of 120µg/L (sensitivity 100.0%; specificity 72.0%). This could be due to participants being at the early stage of CKD. The cut-off values for NLR were a slightly decreased than reports of Singh et al, (24) with cut-off level of 3.28 (sensitivity 89.7%, specificity 69.7%). On the other hand, the cut-off value for NLR in this study is slightly higher than findings of Wan et al, (18) with cut point of 1.7 (sensitivity 68.0, specificity 47.8). These slight variations could be due to ethnic or geographical differences between the populations of study. These findings from the present study, however, suggest that chronic inflammation is involved in development of kidney disease in Type 2 diabetics.

The strength of this study is that, it is the first attempt to investigate the diagnostic value of sNGAL and NLR for the detection of nephropathy in Ghanaian type 2 diabetes mellitus patients. Despite the novel findings, this study has certain limitation. This study was a one-centre study and may not be true reflection of the entire Ghanaian population.

## Conclusion

In conclusion, sNGAL, as an independent biomarker, is an excellent indicator of kidney disease as levels increases in both good and poor glycaemic controlled type 2 diabetics with reduced eGFR. NLR can serve a less expensive and readily measurable alternative biomarker for diabetic nephropathy, particularly in poorly controlled diabetes. These two markers can be added to the available array of tests used to indicate nephropathy in type 2 diabetics to help clinicians in better management of the disease.

## Data Availability

All data produced in the present work are contained in the manuscript

## Data Availability Statement

The data that support the findings of this study are available from the corresponding authors upon reasonable request.

## Declarations

This study was conducted based on the Helsinki Declaration and the study protocol, consent forms and participant information material were reviewed and approved by the Committee for Human Research Publication and Ethics of Kwame Nkrumah University of Science and Technology, School of Medical Sciences (CHRPE/AP/742/22). Permission was sought from the research facility of Ejisu Government Hospital before commencement of study. Written consent of individual participants was sought after the aims, benefits, risk and right of withdrawal at any time from the study were well explained to the study participants in English and in the local dialect (mostly Twi) and their consent was obtained.

## Conflicts of Interest

The authors declare that there is no conflict of interest regarding the publication of this article.

## Authors Contribution

**Conceptualization** Allwell Adofo Ayirebi, Benedict Sackey, Enoch Odame Anto, Lilian Antwi-Boateng, Evans Adu Asamoah, Christian Obirikorang and Otchere Addai-Mensah

**Data curation:** Allwell Adofo Ayirebi, Benedict Sackey, Enoch Odame Anto Prince Adoba, Daniel Nii Martey Antonio, Isaac Opoku Antwi, John Agyemang Sah, Stephen Twumasi Wina Ivy Ofori Boadu Patrick Adu, Joachim Baba Domosie, Afua Marfowaa and Joseph Boachie

**Formal analysis:** Stephen Twumasi, Abiba Khalifah, Ebenezer Senu, Joseph Frimpong, Emmanuel Ekow Korsah, Ezekial Ansah, Eugene Arele

**Investigation:** Allwell Adofo Ayirebi, Daniel Nii Martey Antonio, Stephen Twumasi

**Methodology:** Allwell Adofo Ayirebi, Daniel Nii Martey Antonio, Stephen Twumasi

**Project administration:** Allwell Adofo Ayirebi, Benedict Sackey, Enoch Odame Anto

**Resources:** Allwell Adofo Ayirebi

**Software:** Allwell Adofo Ayirebi, Stephen Twumasi

**Supervision:** Allwell Adofo Ayirebi, Wina Ivy Ofori Boadu, Enoch Odame Anto

**Validation:** Enoch Odame Anto, Wina Ivy Ofori Boadu

**Visualization**: Allwell Adofo Ayirebi, Benedict Sackey, Enoch Odame Anto

**Writing-original drtaft:** Allwell Adofo Ayirebi, Stephen Twumasi, Enoch Odame Anto

**Writing-review & editing:** Allwell Adofo Ayirebi, Wina Ivy Ofori Boadu, Benedict Sackey, Lilian Antwi-Boateng, Prince Adoba, Patrick Adu, Joseph Boachie, Stephen Twumasi, Joseph Frimpong, Emmanuel Ekow Korsah, Ezekial Ansah, Ebenezer Senu, Eugene Arele Ansah, Daniel Nii Martey Antonio, Afua Marfowaa, Isaac Opoku Antwi, John Agyemang Sah, Joachim Baba Domosie, Abiba Khalifah, Evans Adu Asamoah, Christian Obirikorang, Otchere Addai-Mensah, Enoch Odame Anto

## Funding Statement

The authors received no financial support for the research, authorship and or/ publication of this article.

## Acknowledgements

We are grateful for the immense contributions of the staff of Ejisu Government Hospital for their warm reception, not forgetting our participants. Special thanks to the theoretical and applied biology laboratory of Kwame Nkrumah University of Science and Technology for their support in storage of samples.

## Notes

### Competing Interest Statement

The authors have declared no competing interest.

### Funding Statement

This study did not receive any funding

### Author Declarations

The Committee for Human Research Publication and Ethics of Kwame Nkrumah University of Science and Technology, School of Medical Sciences, evaluated and approved the study

## References

1. Guariguata L, Whiting DR, Hambleton I, Beagley J, Linnenkamp U, Shaw JE. Global estimates of diabetes prevalence for 2013 and projections for 2035. Diabetes Res Clin Pract. 2014;103(2). doi 10.1016/j.diabres.2013.11.002

2. Sarfo-Kantanka O, Owusu-Dabo E, Adomako-Boateng F, Eghan B, Dogbe J, Bedu-Addo G. An assessment of prevalence and risk factors for hypertension and diabetes during world diabetes day celebration in Kumasi, Ghana. East Afr J Public Health. 2014;11(2).

3. Asamoah-Boaheng M, Sarfo-Kantanka O, Tuffour AB, Eghan B, Mbanya JC. Prevalence and risk factors for diabetes mellitus among adults in Ghana: A systematic review and meta-analysis. Vol. 11, International Health. 2019. doi 10.1093/inthealth/ihy067

4. Samsu N. Diabetic Nephropathy: Challenges in Pathogenesis, Diagnosis, and Treatment. Vol. 2021, BioMed Research International. 2021. doi 10.1155/2021/1497449

5. Levey AS, Atkins R, Coresh J, Cohen EP, Collins AJ, Eckardt KU, et al. Chronic kidney disease as a global public health problem: Approaches and initiatives - A position statement from Kidney Disease Improving Global Outcomes. In: Kidney International. 2007. doi 10.1038/sj.ki.5002343

6. Ofori E, Gyan KF, Gyabaah S, Nguah SB, Sarfo FS. Predictors of rapid progression of estimated glomerular filtration rate among persons living with diabetes and/or hypertension in Ghana: Findings from a multicentre study. J Clin Hypertens. 2022;24(10). doi 10.1111/jch.14568

7. Okyere P, Okyere I, Ephraim RKD, Attakorah J, Osafo C, Arhin B, et al. Spectrum and Clinical Characteristics of Renal Diseases in Ghanaian Adults: A 13-Year Retrospective Study. Int J Nephrol. 2020;2020. doi 10.1155/2020/8967258

8. Satirapoj B, Nast CC, Adler SG. Novel Insights into the Relationship Between Glomerular Pathology and Progressive Kidney Disease. Vol. 19, Advances in Chronic Kidney Disease. 2012. doi 10.1053/j.ackd.2011.12.001

9. Satirapoj B. Tubulointerstitial Biomarkers for Diabetic Nephropathy. Vol. 2018, Journal of Diabetes Research. 2018. doi 10.1155/2018/2852398

10. Gad M, Kazibwe J, Abassah-Konadu E, Amankwah I, Owusu R, Gulbi G, et al. The Epidemiological and Economic Burden of Diabetes in Ghana: A Scoping Review to Inform Health Technology Assessment. medRxiv. 2023 Jan;2023.04.19.23288806. doi 10.1101/2023.04.19.23288806

11. Bolignano D, Coppolino G, Romeo A, De Paola L, Buemi A, Lacquaniti A, et al. Neutrophil gelatinase-associated lipocalin (NGAL) reflects iron status in haemodialysis patients. Nephrol Dial Transplant. 2009;24(11). doi 10.1093/ndt/gfp310

12. Cernaro V, Bolignano D, Buemi A, Lacquaniti A, Santoro D, Buemi M. Biomarkers in Kidney Disease. Biomarkers Kidney Dis. 2016;1–24. doi 10.1007/978-94-007-7743-9

13. Zhang J, Zhang R, Wang Y, Wu Y, Li H, Han Q, et al. Effects of neutrophil–lymphocyte ratio on renal function and histologic lesions in patients with diabetic nephropathy. Nephrology. 2019;24(11). doi 10.1111/nep.13517

14. Book Reviews. Community Health Stud. 2010;11(2). doi 10.1111/j.1753-6405.1987.tb00145.x

15. Tannor EK, Sarfo FS, Mobula LM, Sarfo-Kantanka O, Adu-Gyamfi R, Plange-Rhule J. Prevalence and predictors of chronic kidney disease among Ghanaian patients with hypertension and diabetes mellitus: A multicenter cross-sectional study. J Clin Hypertens. 2019;21(10). doi 10.1111/jch.13672

16. Levey AS, Stevens LA. Estimating GFR Using the CKD Epidemiology Collaboration (CKD-EPI) Creatinine Equation: More Accurate GFR Estimates, Lower CKD Prevalence Estimates, and Better Risk Predictions. Vol. 55, American Journal of Kidney Diseases. 2010. doi 10.1053/j.ajkd.2010.02.337

17. Paulus I, Palguna KAS, Wirasugianto J, Supadmanaba IG, Semadi IMS, Suastika K. Neutrophil Lymphocyte Ratio (NLR) was Significantly Associated with Diabetic Nephropathy at Sanglah General Hospital, Denpasar, Bali, Indonesia: A Case-Control Study. World J Curr Med Pharm Res. 2021; doi 10.37022/wjcmpr.vi.173

18. Wan H, Wang Y, Fang S, Chen Y, Zhang W, Xia F, et al. Associations between the Neutrophil-to-Lymphocyte Ratio and Diabetic Complications in Adults with Diabetes: A Cross-Sectional Study. J Diabetes Res. 2020;2020. doi 10.1155/2020/6219545

19. Hussain M, Babar MZM, Akhtar L, Hussain MS. Neutrophil lymphocyte ratio (NLR): A well assessment tool of glycemic control in Type-2 diabetic patients. Pakistan J Med Sci. 2017;33(6). doi 10.12669/pjms.336.12900

20. Kaul A, Behera M, Rai M, Mishra P, Bhaduaria D, Yadav S, et al. Neutrophil gelatinase-Associated lipocalin: As a predictor of early diabetic nephropathy in Type 2 diabetes mellitus. Indian J Nephrol. 2018;28(1). doi 10.4103/ijn.IJN_96_17

21. Bolignano D, Lacquaniti A, Coppolino G, Donato V, Campo S, Fazio MR, et al. Neutrophil gelatinase-associated lipocalin (NGAL) and progression of chronic kidney disease. Clin J Am Soc Nephrol. 2009;4(2). doi 10.2215/CJN.03530708

22. Danquah M, Owiredu WKBA, Jnr BAE, Serwaa D, Odame Anto E, Peprah MO, et al. Diagnostic value of neutrophil gelatinase-associated lipocalin (NGAL) as an early biomarker for detection of renal failure in hypertensives: a case–control study in a regional hospital in Ghana. BMC Nephrol [Internet]. 2023;24(1):114. Available from: 10.1186/s12882-023-03120-6 doi 10.1186/s12882-023-03120-6

23. Bolignano D, Coppolino G, Campo S, Aloisi C, Nicocia G, Frisina N, et al. Urinary Neutrophil Gelatinase-Associated Lipocalin (NGAL) is associated with severity of renal disease in proteinuric patients. Vol. 23, Nephrology Dialysis Transplantation. 2008. doi 10.1093/ndt/gfm541

24. Singh A, Jha AK, Kalita BC, Jha DK, Alok Y. Neutrophil lymphocyte ratio: a reliable biomarker for diabetic nephropathy? Int J Diabetes Dev Ctries. 2021; doi 10.1007/s13410-021-01000-z

